# Beyond the Index: Association of AI-CO Central Image Area with Central Visual Field Damage in Glaucoma

**DOI:** 10.64898/2026.10.18.26365818

**Authors:** Yukiko Kora, Hiromi Kora, Toshitake Kora

## Abstract

**Purpose:** To evaluate the associations of the numerical AI-CO center index and visually classified central dark-area extent with Humphrey 10-2 mean deviation (MD).

**Methods:** This single-center retrospective study included 320 eyes of 169 patients with AI-CO imaging, broad-field 24-2F or 30-2 testing, and 10-2 testing. Both visual field examinations were performed within 365 days before or after AI-CO imaging. Two ophthalmologists independently classified central dark-area extent into five Levels (0–4). Analyses assessed Spearman correlations, areas under the receiver operating characteristic curve (AUCs), distributions across final central dark-area Levels, and interrater reliability. Patient-cluster bootstrap confidence intervals and a one-eye-per-patient sensitivity analysis addressed inclusion of both eyes.

**Results:** AI-CO center correlated with 10-2 MD (ρ=0.829; 95% CI, 0.773–0.873). AUCs for 10-2 MD below −2, −6, and −12 dB were 0.902, 0.939, and 0.961, respectively. The numbers of eyes in Levels 0–4 were 66, 57, 94, 53, and 50; median 10-2 MD values were −0.59, −1.48, −3.54, −6.24, and −18.15 dB, respectively. Quadratic-weighted kappa was 0.970 (95% CI, 0.958–0.981). Sensitivity analyses yielded broadly similar findings.

**Conclusions:** The numerical AI-CO center index and visually classified central dark-area extent were associated with central visual field damage. Assessing the extent of dark regions in the central image alongside the numerical index may provide useful information for interpreting central visual field status from the AI-CO display.

## Introduction

Visual field testing is essential in glaucoma care to characterize the severity and distribution of functional damage, assess progression, and guide treatment. Static perimetry quantitatively measures sensitivity at predefined test locations, but the area examined and the density of test locations differ among programs. The 10-2 program, with its dense sampling of the central visual field, is useful for detecting central visual field damage that may not be captured by 24-2 testing [1,2]. Nevertheless, 10-2 testing, which assesses the central visual field relevant to daily functioning, may be underused in routine care [1]. Adding 10-2 testing to broader programs such as 24-2 or 30-2 requires additional time and patient concentration. Clinicians must therefore decide which eyes should undergo additional central testing.

With the widespread use of optical coherence tomography (OCT), methods for estimating visual field damage from retinal structural information have been investigated. Deep learning models have been developed to estimate pointwise visual field sensitivity and global visual field indices from retinal thickness maps or unsegmented OCT images [3,4]. Research models have also been investigated for estimating the central 10-degree visual field from OCT images; some incorporate 24-2 or 30-2 visual field results as additional inputs [5–7]. Koyama et al. reported a research model that estimates 24-2 and 10-2 visual fields from OCT images using a segmentation-free three-dimensional convolutional neural network [8]. Although these studies demonstrate technical feasibility, the meaning and limitations of outputs from commercially implemented systems require evaluation in independent clinical populations.

AI-CO (DeepEyeVision Co., Ltd., Japan) is medical device software that processes three-dimensional OCT image data and presents two-dimensional grayscale and pattern-deviation-like images as reference information related to visual function. The software has received medical device certification in Japan, and the manufacturer announced nationwide commercial sales in January 2026 [9]. In addition to numerical indices, AI-CO displays images that allow visual assessment of the location and extent of the areas shown as abnormal. However, publicly available information on the internal model, training data, and methods used to calculate and calibrate its numerical indices is limited. Researchers therefore cannot independently establish the correspondence between previously published research models and the commercial product.

Humphrey mean deviation (MD) summarizes sensitivity loss across the tested field, whereas image area represents the extent of an abnormal region; these measures therefore contain different information. Similarly, although AI-CO numerical indices summarize image information, the displayed images may contain spatial information not fully represented by the numerical values alone. Examining how the extent of dark areas in the AI-CO central image relates to measured central visual field damage may help clinicians interpret the image display. Examining central image findings in eyes with relatively preserved broad-field 24-2 or 30-2 MD is also relevant to assessing their potential as a cue to consider 10-2 testing.

In a previous study, we evaluated the association between AI-CO all, the broad-field numerical index provided by AI-CO, and Humphrey 24-2 or 30-2 MD in routine clinical care [10]. The present study focused on the central visual field. In addition to evaluating the association between AI-CO center and Humphrey 10-2 MD, we classified dark regions in the AI-CO central image into central dark-area Levels and examined their relationships with the numerical index and 10-2 MD. We also examined the distribution of central dark-area Levels across broad-field and 10-2 MD severity categories to explore the clinical meaning of the image area information.

## Methods

### Study design and participants

This retrospective, single-center observational study evaluated the central output of a commercially implemented artificial intelligence system in patients receiving care for glaucoma at Kora Eye Clinic, Shizuoka, Japan. The study was conducted in accordance with the Declaration of Helsinki and approved by the Institutional Review Board of Jichi Medical University Hospital (approval No. Rindai 26-068; August 21, 2026). Study information was publicly disclosed, and patients were given the opportunity to opt out.

The cohort was drawn from the 819-eye population evaluated in our companion study of the association between AI-CO all and Humphrey 24-2F or 30-2 mean deviation (MD) [10]. For the present study, we included eyes with a corresponding Humphrey 24-2 SITA Faster (24-2F) or 30-2 SITA Fast examination, and a Humphrey 10-2 examination. Both the broad-field and 10-2 examinations were required to be within 365 days before or after AI-CO imaging. When multiple eligible visual field examinations were available, the examination closest in date to AI-CO imaging was selected separately for the broad-field and 10-2 assessments. The selected broad-field examination determined assignment to the 24-2F or 30-2 group. Central dark-area Levels were classified for all included eyes. The final cohort comprised 320 eyes of 169 patients. AI-CO imaging dates ranged from March 31 to May 8, 2026. Both eyes were included when eligible, with one set of measurements per eye. No additional uniform exclusion criterion based on perimetric reliability indices or ocular comorbidity was applied.

### Visual field testing and OCT imaging

Broad-field visual field examinations were performed using the Humphrey Field Analyzer II-i or Humphrey Field Analyzer 3 model 840 (Carl Zeiss Meditec). The 24-2F group comprised eyes examined using the 24-2 program with SITA Faster on the HFA3 840. The 30-2 group comprised eyes examined using the 30-2 program with SITA Fast on either perimeter. Central visual field examinations were performed using the Humphrey 10-2 program with SITA Fast throughout the study. Central visual field status was assessed using the 10-2 MD in both groups. Thus, the program groups refer to the accompanying broad-field examination, whereas the central functional reference was the 10-2 examination throughout. OCT images were obtained with a Topcon TRITON system using the 12 × 9-mm wide-scan mode. For broad-field and 10-2 testing separately, the examination interval was calculated as the absolute difference in days between AI-CO imaging and the selected visual field examination.

### AI-CO indices

AI-CO (DeepEyeVision Co., Ltd.) processes three-dimensional OCT image data and displays visual-function-related reference information, including grayscale and pattern-deviation-like images. The software provides numerical outputs labeled “Peripheral” and “Central,” operationally termed AI-CO all and AI-CO center, respectively. Both are unitless indices. AI-CO center was the principal index evaluated against 10-2 MD. The exact spatial definition and conversion of the proprietary numerical output are unavailable; the analyses therefore assessed association and discrimination rather than agreement or interchangeability with decibel measurements. No data from Kora Eye Clinic were used to develop, train, or tune the model. The same AI-CO software version was used throughout the imaging period from March 31 to May 8, 2026. A representative AI-CO image and corresponding Humphrey visual field reports are shown in Figure 1.

**Figure 1.**
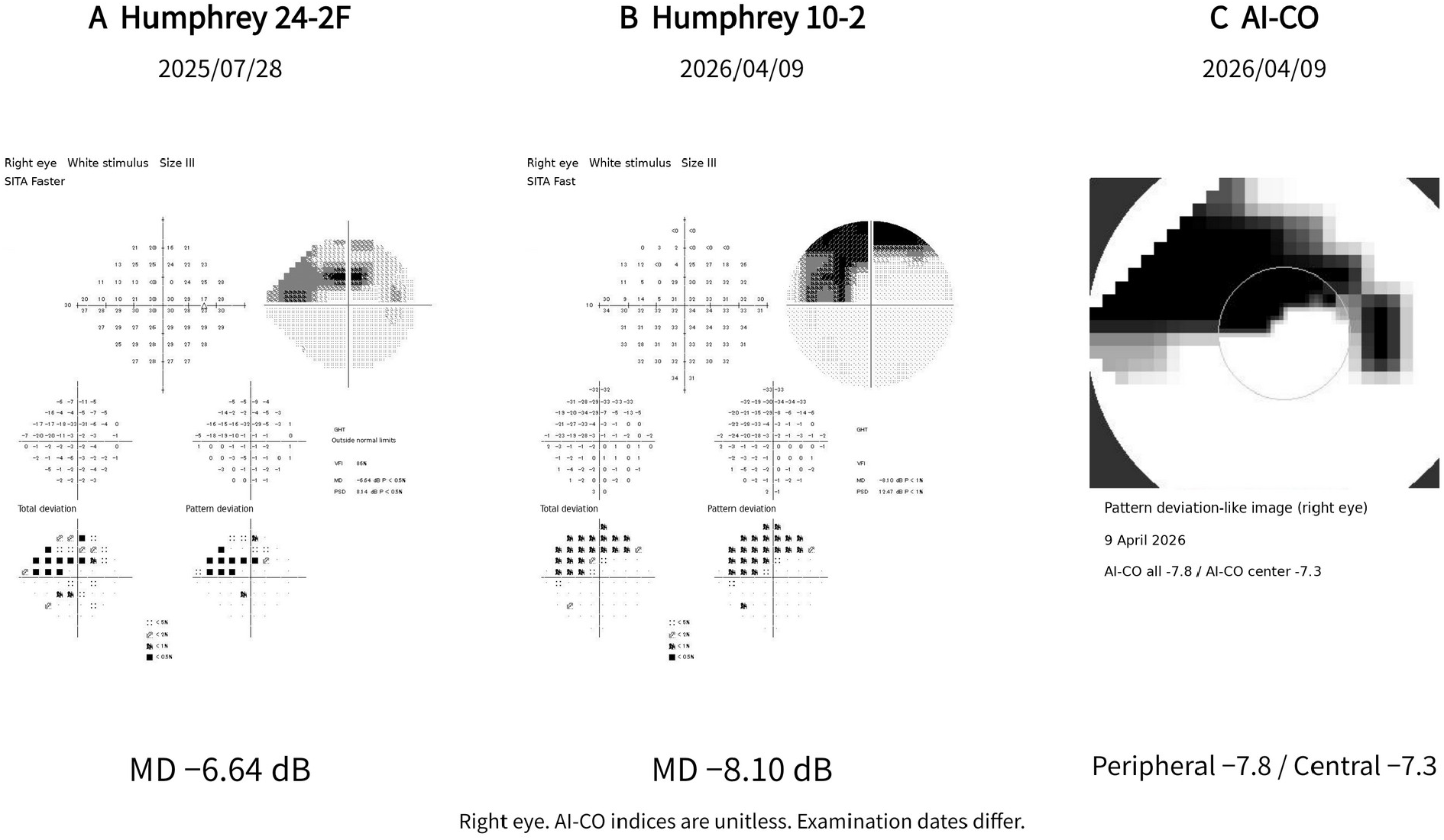
Humphrey visual field reports and the AI-CO image from the same right eye. (A) Humphrey 24-2 SITA Faster examination performed on 28 July 2025 (MD −6.64 dB). (B) Humphrey 10-2 SITA Fast examination performed on 9 April 2026 (MD −8.10 dB). (C) AI-CO pattern-deviation-like image acquired on 9 April 2026; AI-CO all −7.8 and AI-CO center −7.3 are unitless indices. The outlined circle indicates the central region. The Humphrey panels retain the threshold values, grayscale plots, total deviation and pattern deviation values and probability maps from the anonymized source reports. Japanese labels were replaced with English labels, and selected examination metadata were summarized in English. Plotted values and map orientation were preserved. The display terms Peripheral and Central correspond to AI-CO all and AI-CO center, respectively. The image annotation “Examination dates differ” refers to the earlier 24-2F examination; the 10-2 examination and AI-CO imaging were performed on the same day. This example does not establish that AI-CO prompted the 10-2 examination.

### Outcomes and severity definitions

The principal analyses were the Spearman rank correlation between AI-CO center and 10-2 MD and the discrimination of 10-2 MD below −2, −6, and −12 dB. For descriptive analyses, eyes were classified as normal range (MD ≥−2 dB), mild (−6 dB ≤MD <−2 dB), moderate (−12 dB ≤MD <−6 dB), or severe (MD <−12 dB). These MD-only categories were defined for this study and do not represent the complete Hodapp–Parrish– Anderson classification or a diagnostic definition of central visual field damage. Secondary analyses examined robustness across broad-field program groups and in a fixed cohort with one eye per patient. Additional analyses described AI-CO center and 10-2 MD across central dark-area Levels and the relationship of AI-CO indices to broad-field MD.

### Classification of central dark-area extent

Two ophthalmologists independently estimated the proportion of the central assessment region occupied by dark areas in the AI-CO pattern-deviation-like image. The estimated extent was classified as Level 0, no dark area (0%); Level 1, 1–25%; Level 2, 26–50%; Level 3, 51–75%; or Level 4, 76–100%. These categories describe visually estimated image area rather than automated pixel measurements. Independent ratings were made without reference to AI-CO numerical indices or Humphrey broad-field or 10-2 results. For analyses involving central dark-area Levels, the first rater’s classification was used for eyes with discrepant independent ratings, with confirmed image-review corrections incorporated into the final classifications. Interrater reliability was calculated using each rater’s independent ratings. Representative examples of the five central dark-area Levels are shown in Supplementary Figure 1.

### Statistical analysis

Continuous variables were summarized as mean and standard deviation or median and interquartile range (IQR), and categorical variables as counts and percentages. Demographic summaries were calculated at the eye level. Spearman rank correlations were calculated overall and separately for the 24-2F and 30-2 groups. The overall Pearson correlation between AI-CO center and 10-2 MD was calculated as a supplementary descriptive measure of linear association. Supplementary Spearman correlations assessed AI-CO all and AI-CO center against broad-field MD. For eye-level correlation and AUC analyses, 95% confidence intervals (CIs) were estimated using 2,000 patient-cluster bootstrap resamples. Patients were ordered by patient identifier, and each resample selected 169 patients with replacement and retained all eligible eyes of each selected patient, including repeated copies when a patient was selected more than once. The overall and program-specific estimates were calculated from the same resample; program-specific estimates used the eyes belonging to the respective program within that resample. Patients were not resampled separately within program groups. Percentile intervals were taken from the 2.5th and 97.5th percentiles of the bootstrap estimates. These analyses used NumPy default_rng with the PCG64 generator and seed 20261016.

For ROC analyses, eyes with 10-2 MD below each threshold were positive and those at or above the threshold were negative. Because lower AI-CO center values indicate greater impairment, −AI-CO center was used as the classification score. Empirical AUCs incorporated half credit for tied scores. Between-program differences were defined as the 24-2F estimate minus the 30-2 estimate for both correlations and AUCs. Differences were calculated within the same patient-cluster bootstrap resamples and described using percentile 95% CIs. No clinical cutoff was derived in this study.

Sensitivity analyses were repeated in a fixed cohort comprising one eligible eye per patient (169 eyes). One eye per patient was randomly selected before final data verification (NumPy default_rng, PCG64; seed 20260818). The selection was retained for eligible eyes; when a selected eye was excluded, the remaining eligible fellow eye was used. The same final 169-eye selection was used across all sensitivity analyses. Confidence intervals in this cohort were estimated using 2,000 resamples of the 169 patients, ordered by patient identifier, with NumPy default_rng (PCG64) initialized separately with seed 20261016.

Interrater reliability for the ordinal central dark-area Levels was assessed primarily using quadratic-weighted Cohen’s kappa. Linear-weighted and unweighted kappa, exact agreement, and agreement within one Level were also calculated. Quadratic-weighted kappa, exact agreement, and the percentile 95% confidence interval for quadratic-weighted kappa were calculated using an R script (R version 4.5.2) in EZR version 1.70 (Saitama Medical Center, Jichi Medical University, Saitama, Japan) [11]. The confidence interval was estimated using 5,000 patient-cluster bootstrap resamples of the 169 patients with paired ratings, retaining all paired eyes from each sampled patient. Patients were ordered by patient identifier as strings. Resampling used the Mersenne-Twister generator with Inversion and Rejection settings and seed 20261009. Linear-weighted and unweighted kappa and agreement within one Level were calculated from the same paired-rating contingency table using Python with NumPy.

AI-CO center and 10-2 MD distributions across the five central dark-area Levels were compared using Kruskal– Wallis tests. Because these tests in the 320-eye cohort treat eyes as independent, their P values were considered exploratory. The same tests were also performed in the one-eye-per-patient cohort. No formal test of a monotonic trend or pairwise Level differences was performed. Tests were two-sided, with P<0.05 used for exploratory inference. Correlation, ROC, distribution, and non-kappa bootstrap analyses used Python 3.12.14 with NumPy 2.3.5 and SciPy 1.17.0; openpyxl was used to extract the spreadsheet data. Yukiko Kora independently verified Spearman correlation and ROC AUC point estimates for AI-CO center and 10-2 MD and Kruskal–Wallis statistics using EZR. Distributions are shown in Figures 2 and 3. Central dark-area Level distributions were also tabulated within broad-field and central MD categories using the same −2, −6, and −12 dB boundaries (Figures 4 and 5).

**Figure 2.**
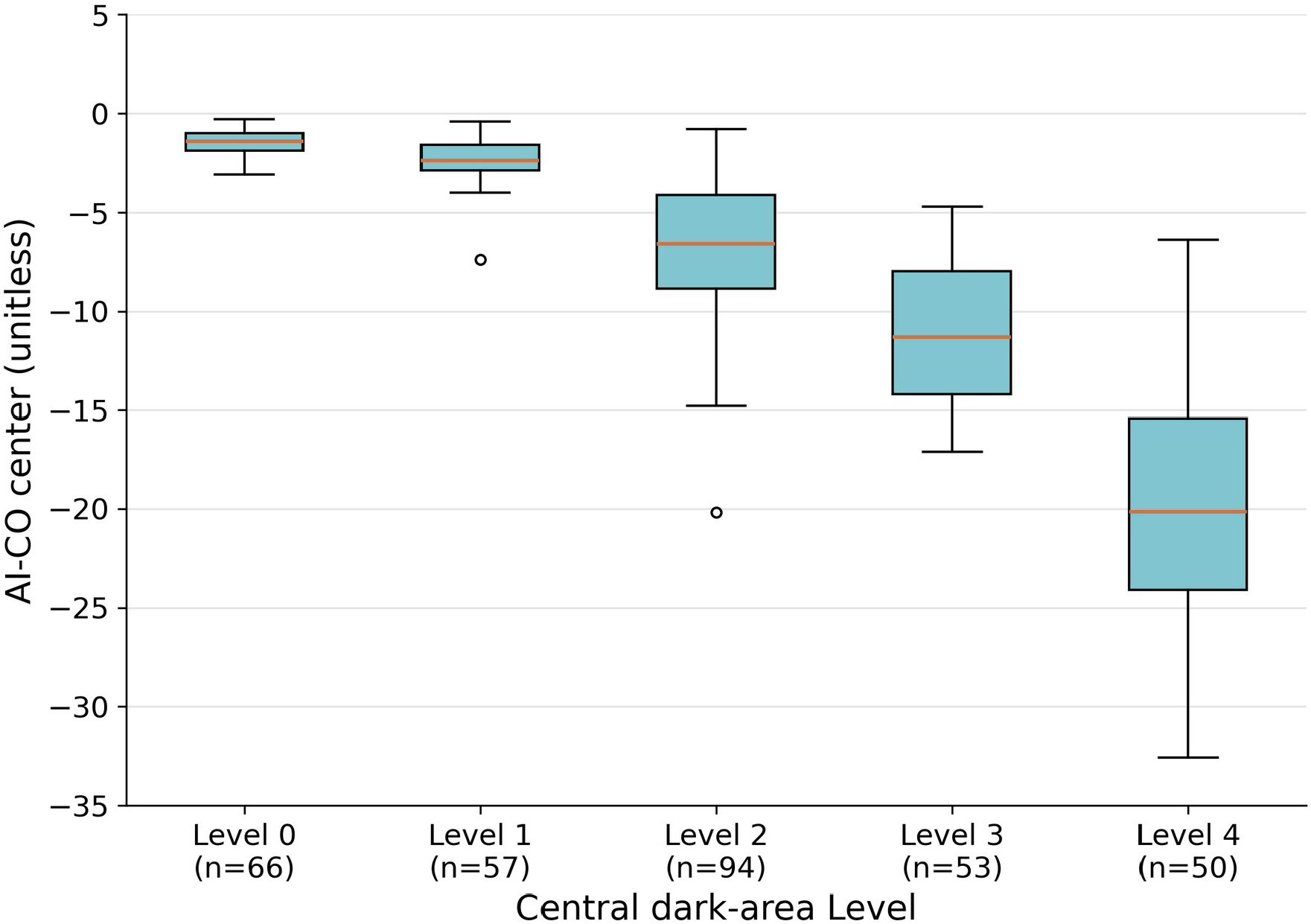
Distribution of AI-CO center by central dark-area Level. Boxes show the interquartile range, horizontal lines indicate medians, whiskers extend to the most extreme values within 1.5 times the interquartile range, and circles indicate values beyond the whiskers. All 320 eyes are included. AI-CO center is unitless; Humphrey MD is measured in dB. The exploratory eye-level Kruskal–Wallis test gave P<0.001. Level 0 denotes no dark area (0%); Levels 1–4 denote 1–25%, 26–50%, 51–75%, and 76–100% of the central assessment region, respectively.

**Figure 3.**
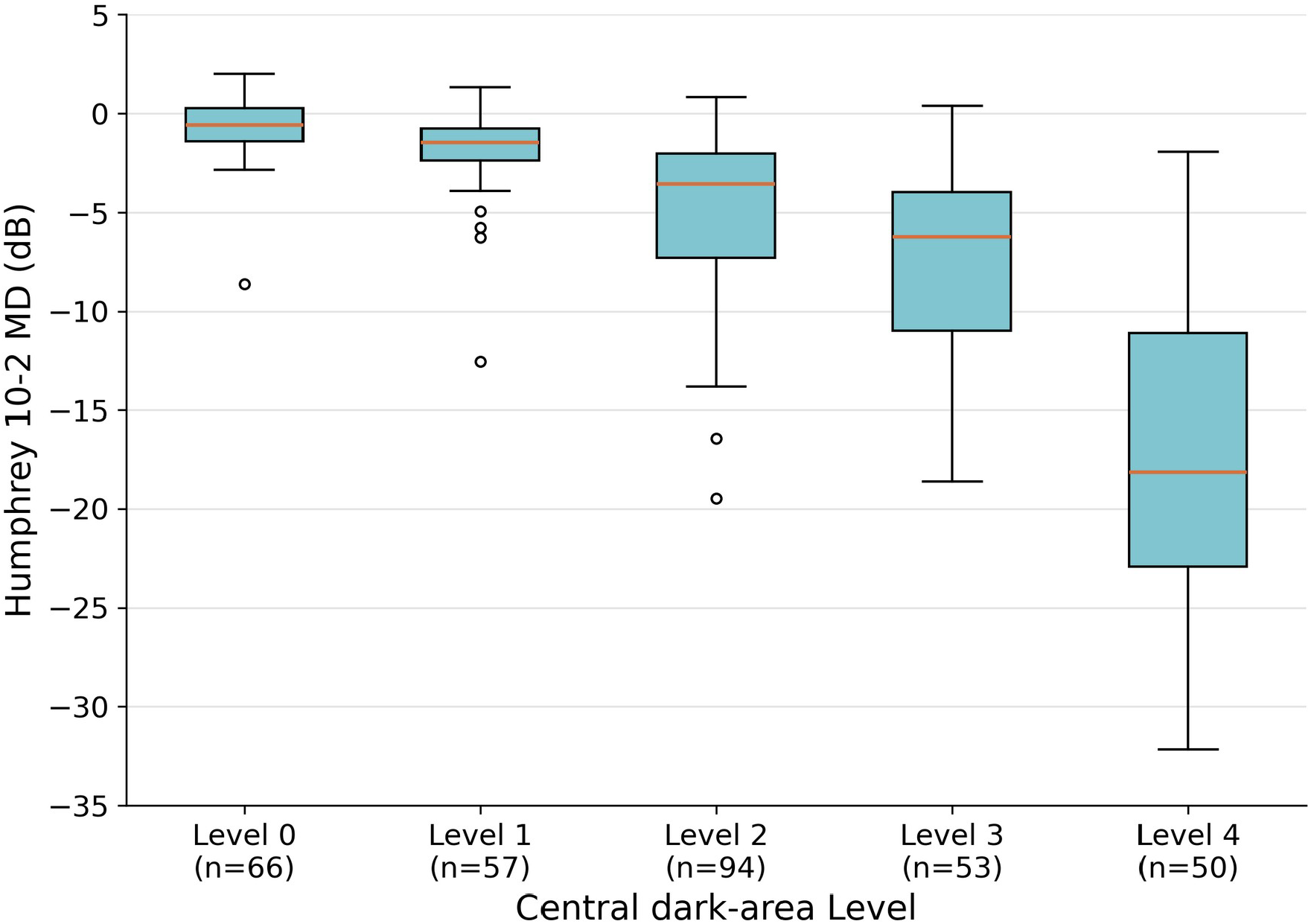
Distribution of Humphrey 10-2 MD by central dark-area Level. Boxes show the interquartile range, horizontal lines indicate medians, whiskers extend to the most extreme values within 1.5 times the interquartile range, and circles indicate values beyond the whiskers. All 320 eyes are included. AI-CO center is unitless; Humphrey MD is measured in dB. The exploratory eye-level Kruskal–Wallis test gave P<0.001. Level 0 denotes no dark area (0%); Levels 1–4 denote 1–25%, 26–50%, 51–75%, and 76–100% of the central assessment region, respectively.

**Figure 4.**
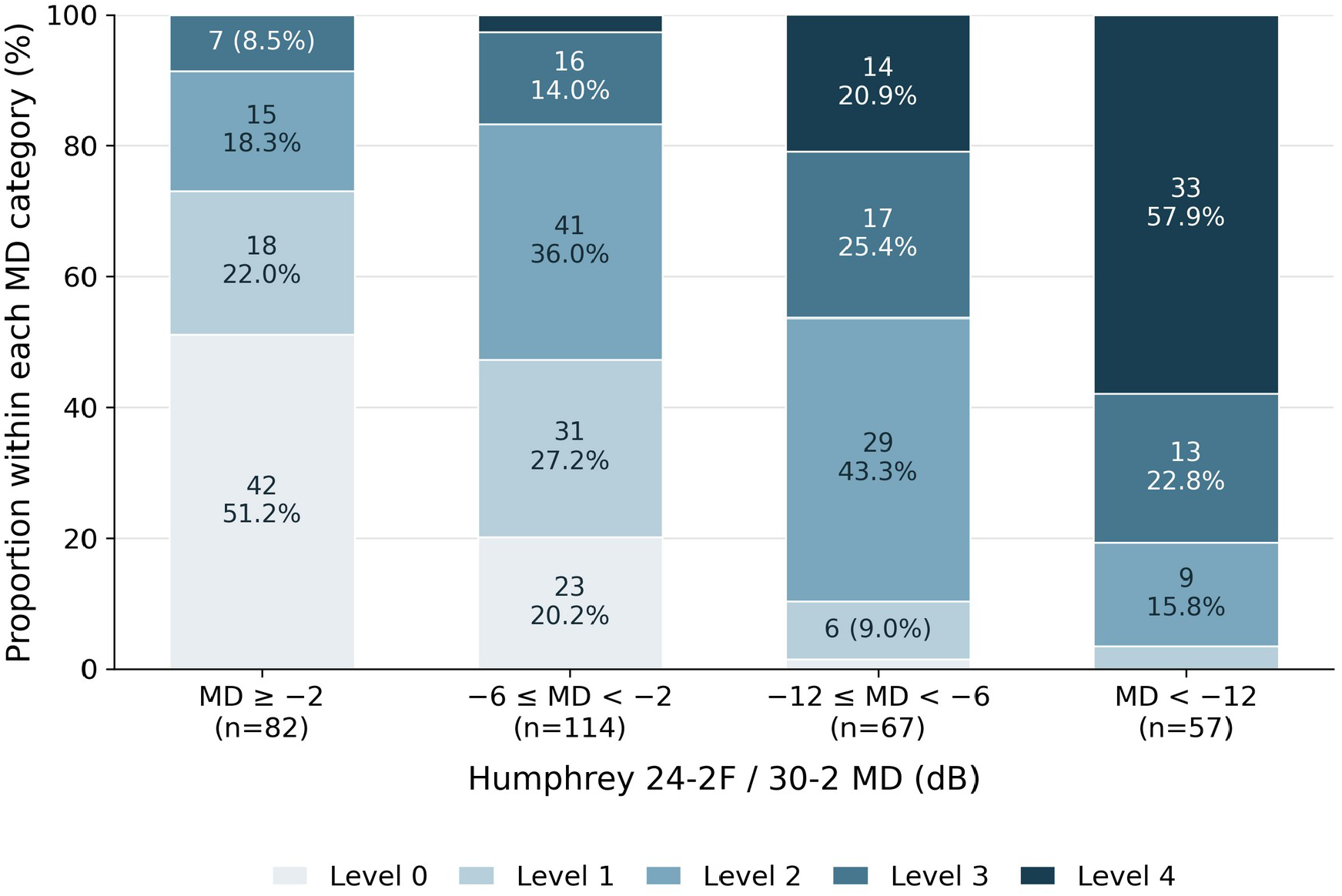
Central dark-area Level distribution across broad-field MD categories. Categories were defined using the MD from the accompanying Humphrey 24-2F or 30-2 examination. The four categories are MD ≥−2 dB, −6≤MD <−2 dB, −12≤MD <−6 dB, and MD <−12 dB. Stacked bars show the percentage of eyes in each Level within each MD category; labels show eye counts and percentages for segments of at least 7%. All segments, including smaller unlabeled segments, are included. Category totals are shown below the bars. All 320 eyes are included. These MD-only categories were defined for this study and do not represent the full Hodapp–Parrish– Anderson classification.

**Figure 5.**
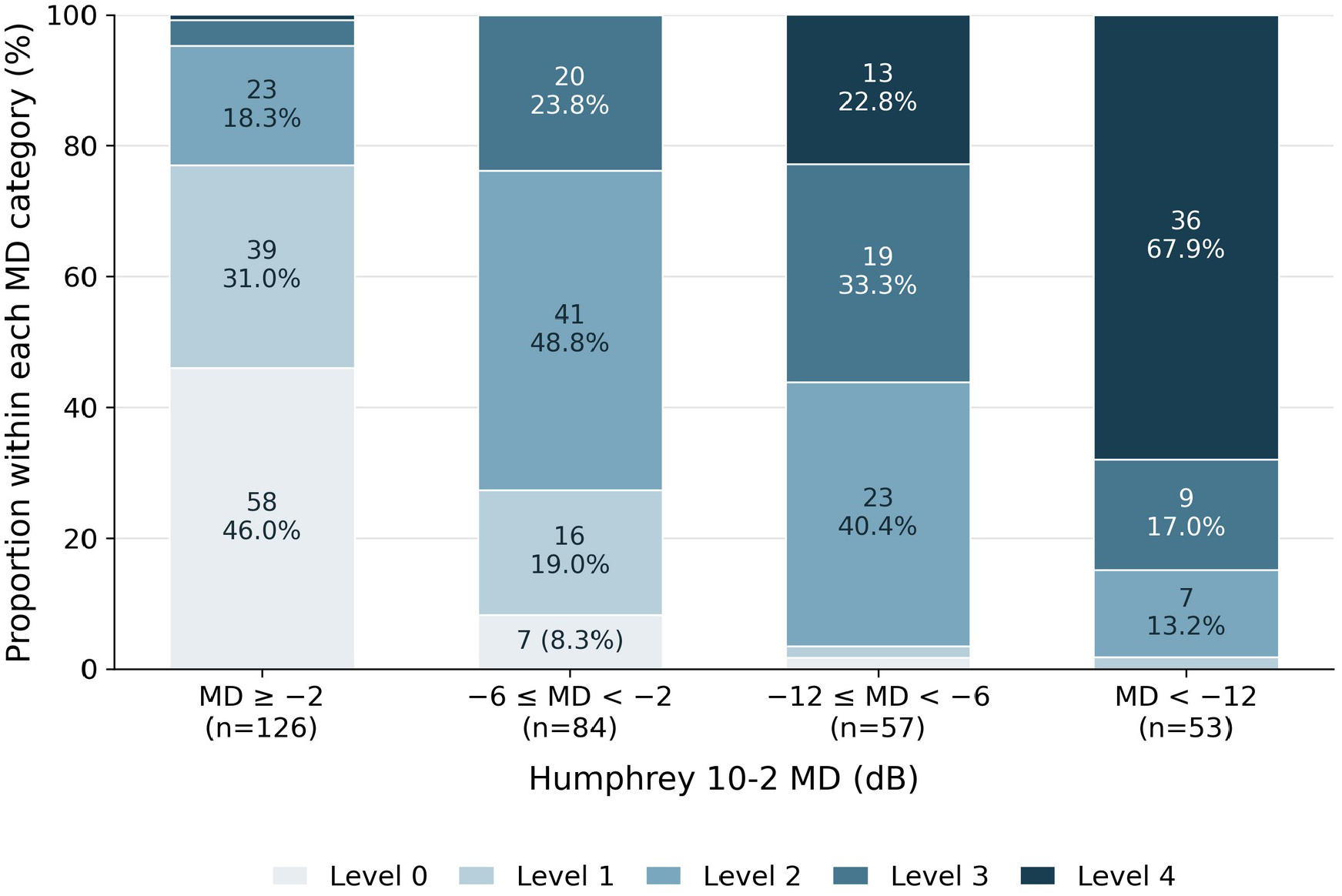
Central dark-area Level distribution across central MD categories. Categories were defined using Humphrey 10-2 MD. The four categories are MD ≥−2 dB, −6≤MD <−2 dB, −12≤MD <−6 dB, and MD <−12 dB. Stacked bars show the percentage of eyes in each Level within each MD category; labels show eye counts and percentages for segments of at least 7%. All segments, including smaller unlabeled segments, are included. Category totals are shown below the bars. All 320 eyes are included. These MD-only categories were defined for this study and do not represent the full Hodapp–Parrish–Anderson classification.

### Declaration of generative AI and AI-assisted technologies

During the preparation of this manuscript in September and October 2026, the authors used GPT-5.6 Sol and GPT-6 through ChatGPT/Codex (OpenAI, https://chatgpt.com) to assist with statistical coding, verification of analytical procedures and numerical consistency, generation of data visualizations, organization and drafting of manuscript content, and English-language editing. Claude Sonnet 5 and Claude Opus 5.5 through Claude (Anthropic, https://claude.ai) were used to review the manuscript for internal consistency and clarity. All analytical methods, interpretations, and final editorial decisions were made and approved by the authors. The authors reviewed the analyses, figures, references, interpretations, and manuscript text and take responsibility for the submitted work.

## Results

### Participants

The final analysis included 320 eyes of 169 patients, comprising 151 patients with both eyes and 18 with one eye. There were 197 eyes in the 24-2F group and 123 in the 30-2 group. The eye-level mean age was 68.6 ± 11.5 years (range, 35–93), and 142 eyes (44.4%) were from male patients. Median 10-2 MD was −3.07 dB (IQR, −8.87 to −1.21), and median AI-CO center was −5.25 (IQR, −11.43 to −2.20). The median absolute interval from AI-CO imaging was 98 days (IQR, 0–190) for broad-field testing and 110.5 days (IQR, 0–190) for 10-2 testing (Table 1). The numbers in the normal-range, mild, moderate, and severe 10-2 MD categories were 126, 84, 57, and 53, respectively.

**Table 1.**
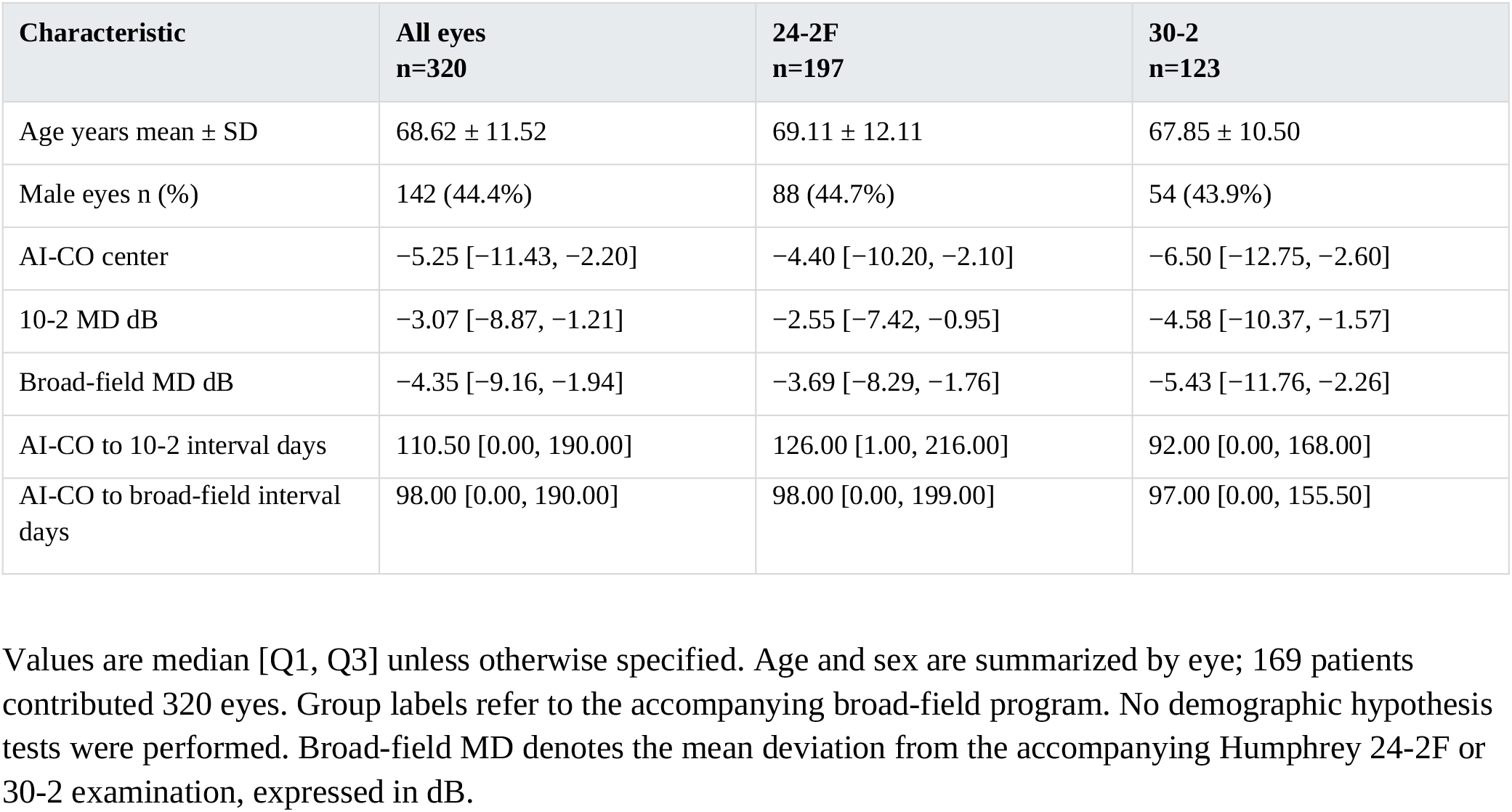
Participant and examination characteristics.

### Association with central visual field severity

AI-CO center was positively associated with 10-2 MD (Spearman ρ=0.829 (0.773–0.873); values in parentheses are patient-cluster 95% CIs). The corresponding correlations were 0.817 (0.736–0.876) in the 24-2F group and 0.847 (0.762–0.907) in the 30-2 group (Table 2). The between-program difference (24-2F minus 30-2) was −0.030 (95% CI, −0.130 to 0.076). These estimates did not establish equivalence between programs. As a supplementary descriptive measure, the overall Pearson correlation was 0.913.

**Table 2.** Spearman correlation between AI-CO center and 10-2 MD.

| Cohort | Eyes | Spearman $\rho$ | 95% CI |
| --- | --- | --- | --- |
| All eyes | 320 | 0.829 | 0.773–0.873 |
| 24-2F | 197 | 0.817 | 0.736–0.876 |
| 30-2 | 123 | 0.847 | 0.762–0.907 |
| One eye per patient | 169 | 0.807 | 0.733–0.866 |

Median AI-CO center values in the normal-range, mild, moderate, and severe 10-2 MD categories were −2.00, −5.05, −10.80, and −20.00, respectively. The corresponding IQRs were −3.05 to −1.20, −7.40 to −3.00, −13.80 to −8.80, and −24.10 to −15.50 (Table 3).

**Table 3.** AI-CO center by central MD severity.

| Category | 10-2 MD dB | Eyes | AI-CO center<br>median [Q1, Q3] |
| --- | --- | --- | --- |
| Normal range | MD ≥ -2 | 126 | -2.00 [-3.05, -1.20] |
| Mild | -6 ≤ MD $<$ -2 | 84 | -5.05 [-7.40, -3.00] |
| Moderate | -12 ≤ MD $<$ -6 | 57 | -10.80 [-13.80, -8.80] |
| Severe | MD $<$ -12 | 53 | -20.00 [-24.10, -15.50] |

### Discrimination of central MD thresholds

AI-CO center discriminated eyes across all three 10-2 MD thresholds. The AUCs were 0.902 (0.866–0.934) for MD <−2 dB, 0.939 (0.905–0.968) for MD <−6 dB, and 0.961 (0.919–0.988) for MD <−12 dB. The numbers of positive eyes were 194, 110, and 53, respectively (Table 4). When eyes were grouped by the accompanying broad-field test (24-2F or 30-2), AUC point estimates ranged from 0.898 to 0.975. The 95% CIs for AUC differences between the 24-2F and 30-2 groups included zero at all three thresholds, indicating no clear evidence of a difference in discrimination between these groups. However, this finding does not establish equivalence (Supplementary Table S1).

**Table 4.** Discrimination of central MD thresholds.

| Positive outcome | Positive eyes | AUC | Patient-cluster 95% CI |
| --- | --- | --- | --- |
| 10-2 MD <-2 dB | 194 | 0.902 | 0.866–0.934 |
| 10-2 MD <-6 dB | 110 | 0.939 | 0.905–0.968 |
| 10-2 MD <-12 dB | 53 | 0.961 | 0.919–0.988 |

### One eye per patient sensitivity analysis

In the fixed 169-eye cohort, the Spearman correlation was 0.807 (0.733–0.866). AUCs were 0.907 (0.861– 0.946), 0.926 (0.876–0.968), and 0.958 (0.896–0.994) at the −2, −6, and −12 dB thresholds, respectively. These estimates were broadly consistent with the 320-eye analysis (Table 2 and Supplementary Table S1).

### Central dark-area Levels

The numbers of eyes in the final classifications for Levels 0–4 were 66, 57, 94, 53, and 50, respectively. Median AI-CO center values were −1.40, −2.40, −6.60, −11.30, and −20.15. Median 10-2 MD values were −0.59, −1.48, −3.54, −6.24, and −18.15 dB (Table 5). Both measures differed across Levels in the exploratory eye-level Kruskal–Wallis tests (AI-CO center, H=256.97; 10-2 MD, H=195.13; both P<0.001). The descriptive medians decreased as Level increased. In the one-eye-per-patient cohort, both Kruskal–Wallis tests also yielded P<0.001. Figures 2 and 3 show the distributions by Level.

**Table 5.**
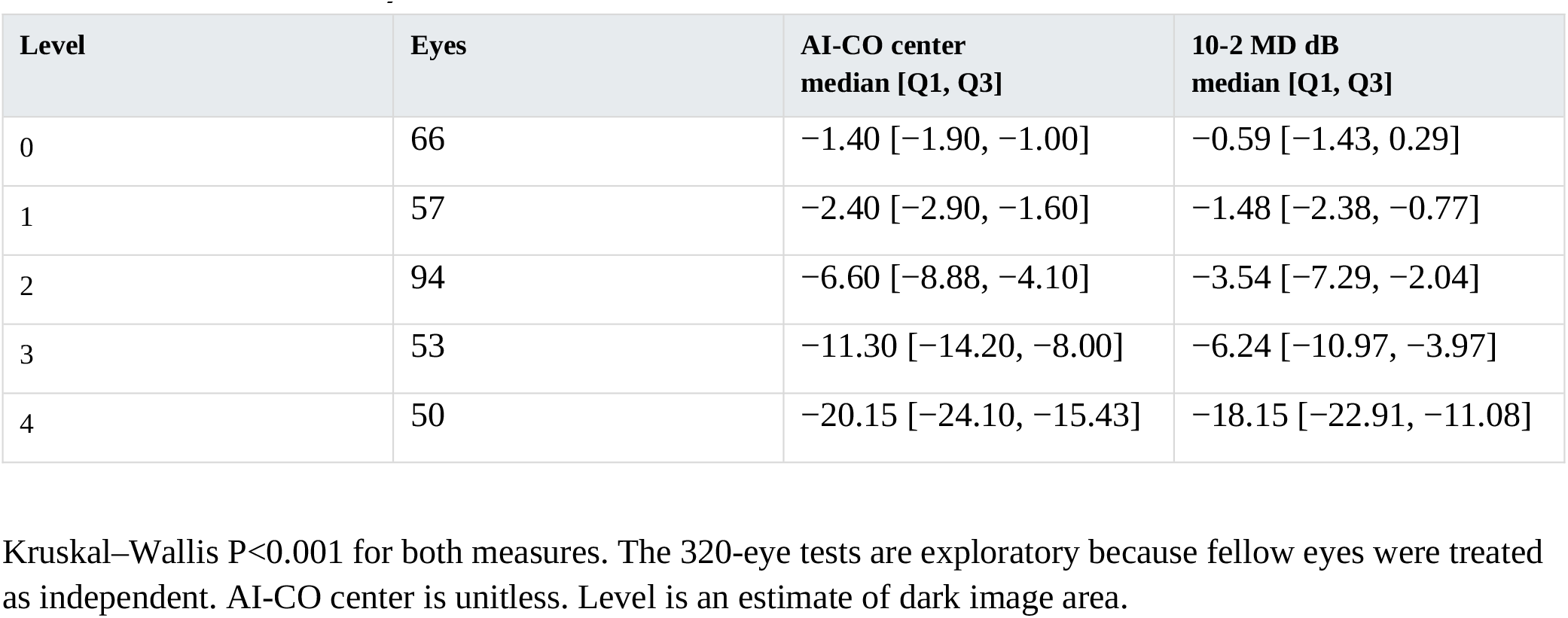
Distributions by central dark-area Level.

### Interrater reliability

Interrater reliability was evaluated in 320 eyes of 169 patients. Quadratic-weighted kappa was 0.970 (95% CI, 0.958–0.981). Linear-weighted and unweighted kappa were 0.931 and 0.869, respectively. The two raters agreed exactly in 287 of 320 eyes (89.7%); all paired ratings differed by no more than one Level. Interrater reliability was assessed using the ratings independently assigned by each rater.

### Supplementary broad-field associations

Within this central-testing cohort, the Spearman correlation between AI-CO all and the MD of the accompanying broad-field test (24-2F or 30-2; hereafter, broad-field MD) was 0.819 (95% CI, 0.769–0.864). The correlation between AI-CO center and broad-field MD was 0.732 (95% CI, 0.672–0.782). These supplementary findings describe associations within the present 320-eye cohort. Figures 4 and 5 show the central dark-area Level distributions across broad-field and central MD categories, respectively. Among eyes with broad-field MD ≥−2 dB, 22 of 82 eyes (26.8%) were classified as Level 2 or 3. Among these 22 eyes, the median 10-2 MD was −1.38 dB, and 7 had 10-2 MD below −2 dB. Among eyes with 10-2 MD ≥−2 dB, 28 of 126 eyes (22.2%) were classified as Level 2 or 3.

## Discussion

AI-CO is glaucoma care support software that presents information on visual field damage estimated from OCT images. However, publicly available information on how its numerical indices and images are generated, its development data, and its internal model is limited. We therefore conducted an exploratory assessment of the associations between the product’s numerical and image outputs and Humphrey visual field results to aid their clinical interpretation.

The Spearman rank correlation between AI-CO center and 10-2 MD was 0.829, and AUCs indicated good discrimination across the −2, −6, and −12 dB MD thresholds. Analyses by program group and with one eye per patient yielded broadly similar results. However, the absence of a clear difference between eyes grouped by the accompanying broad-field test (24-2F or 30-2) does not establish equivalence between the groups. These analyses assessed association and discrimination and do not demonstrate that AI-CO center can replace 10-2 MD.

Whereas our previous study [10] addressed the association between the numerical AI-CO all index and broad-field MD, the present study focused on the central visual field and evaluated image interpretation in addition to numerical outputs. Median 10-2 MD and AI-CO center decreased as the visually classified central dark-area Level increased, and agreement between the two ophthalmologists’ classifications was high. These findings suggest that, alongside the numerical indices of commercially implemented AI-CO, the visually apparent extent of dark regions in the central image is also associated with central visual field status.

The central dark-area Levels provide a simple visual classification intended to reflect image interpretation in routine clinical care. Quadratic-weighted kappa was 0.970 and exact agreement was 89.7%, indicating high agreement between the two raters in this study. Nevertheless, AI-CO center and 10-2 MD varied within the same Level, with greater variability at higher Levels. Dark-area extent describes spatial spread but does not directly quantify the depth or location of damage. MD summarizes sensitivity loss across the tested field and contains information different from area. Moreover, because 10-2 testing measures sensitivity at discrete locations, this study demonstrated an association between dark image area and 10-2 MD, rather than agreement with the actual area of visual field damage. The extent to which damage depth and location contribute to this variability requires further investigation.

A clinically relevant finding was the presence of Level 2 or 3 eyes even in categories with relatively preserved broad-field MD. Central visual field abnormalities missed by 24-2 testing have previously been detected with 10-2 testing [2]. However, evidence is insufficient to support routine 10-2 testing in every eye with early glaucoma [12]; given constraints on time and clinical resources, additional testing in every eye may be difficult to implement in routine care. Limited frequencies of visual field testing in clinical care have also been reported [13]. Findings such as reproducible abnormalities at central 24-2 locations or thinning of the macular inner retina are relevant when considering additional testing [12]. AI-CO estimates information about visual field damage from OCT images and presents it as image displays. In this study, the extent of dark regions in its central image was associated with 10-2 MD. These findings suggest that AI-CO central images may help clinicians recognize the spatial extent of central image abnormalities and consider detailed assessment with 10-2 testing. This study did not assess whether image interpretation provides incremental value beyond the AI-CO center index. In Figure 1, superior abnormalities are also evident on 24-2F, but the AI-CO image draws attention to their central extension, which 10-2 depicts using a denser array of test locations. Supplementary Figure 2 illustrates a localized dark region in the AI-CO central image despite relatively preserved MD on both visual field examinations. However, these illustrative cases do not establish the clinical utility of using AI-CO image findings to guide additional 10-2 testing.

This study has limitations. It was a single-center retrospective study restricted to patients who had already undergone 10-2 testing, and caution is needed when applying the findings to the general glaucoma care population. Eyes with coexisting ocular conditions were not excluded; these conditions may have affected both the AI-CO images and the visual field results. AI-CO imaging and visual field testing were not always performed on the same day, and the potential influence of the examination interval should be considered. Area classification relied on visual assessment by two ophthalmologists. Although interrater reliability was high, reproducibility among other raters and institutions remains to be evaluated. In addition, the internal AI-CO algorithm and the precise spatial correspondence of the central assessment region have not been disclosed, limiting our ability to fully explain the meaning of the image findings.

In conclusion, AI-CO center and the extent of dark regions in the central image were associated with central visual field damage. Interpreting numerical indices together with the images may help clinicians understand central visual field status and consider additional perimetry. Their clinical utility requires validation in prospective studies.

## Data availability

The de-identified data that support the findings of this study are available from the corresponding author upon reasonable request, subject to institutional ethical and privacy requirements.

## Acknowledgments

The authors thank the orthoptists at Kora Eye Clinic for their assistance with data collection.

## References

1. Du KH, Kamalipour A, Moghimi S. Central visual field in glaucoma: An updated review. Taiwan J Ophthalmol. 2024;14(3):360–370. doi:10.4103/tjo.TJO-D-24-00042.

2. De Moraes CG, Hood DC, Thenappan A, et al. 24-2 Visual Fields Miss Central Defects Shown on 10-2 Tests in Glaucoma Suspects, Ocular Hypertensives, and Early Glaucoma. Ophthalmology. 2017;124(10):1449–1456. doi:10.1016/j.ophtha.2017.04.021.

3. Park K, Kim J, Lee J. A deep learning approach to predict visual field using optical coherence tomography. PLoS One. 2020;15:e0234902. doi:10.1371/journal.pone.0234902.

4. Hemelings R, et al. Pointwise visual field estimation from optical coherence tomography in glaucoma using deep learning. Transl Vis Sci Technol. 2022;11:22. doi:10.1167/tvst.11.8.22.

5. Moon S, Lee JH, Choi H, Lee SY, Lee J. Deep learning approaches to predict 10-2 visual field from wide-field swept-source optical coherence tomography en face images in glaucoma. Sci Rep. 2022;12:21041. doi:10.1038/s41598-022-25660-x.

6. Asano S, et al. Predicting the central 10 degrees visual field in glaucoma by applying a deep learning algorithm to optical coherence tomography images. Sci Rep. 2021;11:2214. doi:10.1038/s41598-020-79494-6.

7. Hashimoto Y, et al. Predicting 10-2 visual field from optical coherence tomography in glaucoma using deep learning corrected with 24-2/30-2 visual field. Transl Vis Sci Technol. 2021;10:28. doi:10.1167/tvst.10.13.28.

8. Koyama M, Ueno Y, Ito Y, Oshika T, Tanito M. Automated learning of glaucomatous visual fields from OCT images using a comprehensive, segmentation-free 3D convolutional neural network model. Sci Rep. 2025;15:13395. doi:10.1038/s41598-025-98511-0.

9. DeepEyeVision Co., Ltd. Announcement of nationwide commercial sales of AI-CO medical device software. https://deepeyevision.com/news20260122/ (22 January 2026; accessed 7 October 2026).

10. Kora Y, Kora H, Kora T. Real-World External Validation of a Clinically Implemented Artificial Intelligence Index for Visual Field Severity from Optical Coherence Tomography. medRxiv [Preprint]. 2026. doi:10.64898/2026.09.08.26362248.

11. Kanda Y. Investigation of the freely available easy-to-use software ‘EZR’ for medical statistics. Bone Marrow Transplant. 2013;48(3):452–458. doi:10.1038/bmt.2012.244.

12. WuDunn D, Takusagawa HL, Rosdahl JA, et al. Central Visual Field Testing in Early Glaucoma: A Report by the American Academy of Ophthalmology. Ophthalmology. 2024;131(2):240–248. doi:10.1016/j.ophtha.2023.10.008.

13. Stagg BC, et al. The frequency of visual field testing in a US nationwide cohort of individuals with open-angle glaucoma. Ophthalmol Glaucoma. 2022;5:587–593. doi:10.1016/j.ogla.2022.05.002.

