## Supplementary Table S1 and Figures S1-S2 for "Beyond the Index: Association of AI-CO Central Image Area with Central Visual Field Damage in Glaucoma"

### Supplementary Table S1 AUCs by program and sensitivity cohort

| **10-2 threshold** | **24-2F AUC**<br>**95% CI** | **30-2 AUC**<br>**95% CI** | **169-eye AUC**<br>**95% CI** |
|---|---|---|---|
| MD <−2 dB | 0.898 (0.853–0.940) | 0.909 (0.850–0.957) | 0.907 (0.861–0.946) |
| MD <−6 dB | 0.949 (0.903–0.980) | 0.933 (0.878–0.975) | 0.926 (0.876–0.968) |
| MD <−12 dB | 0.975 (0.950–0.992) | 0.944 (0.861–0.997) | 0.958 (0.896–0.994) |

AUC differences (24-2F minus 30-2) were −0.011 (95% CI, −0.077 to 0.062), 0.016 (−0.046 to 0.077), and 0.031 (−0.029 to 0.117), respectively. All intervals use 2,000 patient resamples with NumPy default_rng (PCG64), seed 20261016. Program estimates and between-program differences were calculated from the same full-cohort patient resamples. The one-eye-per-patient analysis used a separately initialized generator with the same seed.

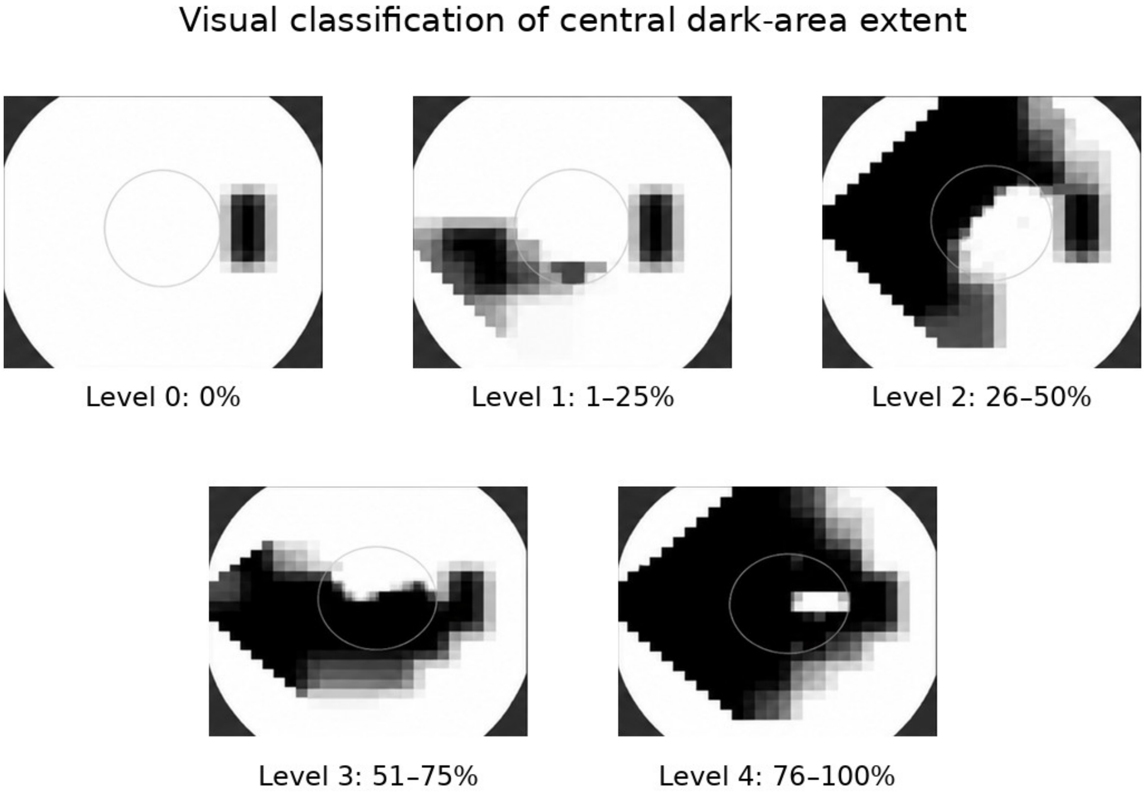


Supplementary Figure 1. Representative examples of the visual classification of central dark-area extent in AI-CO pattern-deviation-like images. The proportion of dark area within the outlined central circle was visually estimated and classified as Level 0, no dark area (0%); Level 1, 1–25%; Level 2, 26–50%; Level 3, 51–75%; or Level 4, 76–100%. These Levels describe visually estimated image area, rather than automated pixel measurements or the depth of visual field loss. The images were cropped from the source display to remove interface labels and numerical indices; image orientation and the displayed dark regions were preserved.

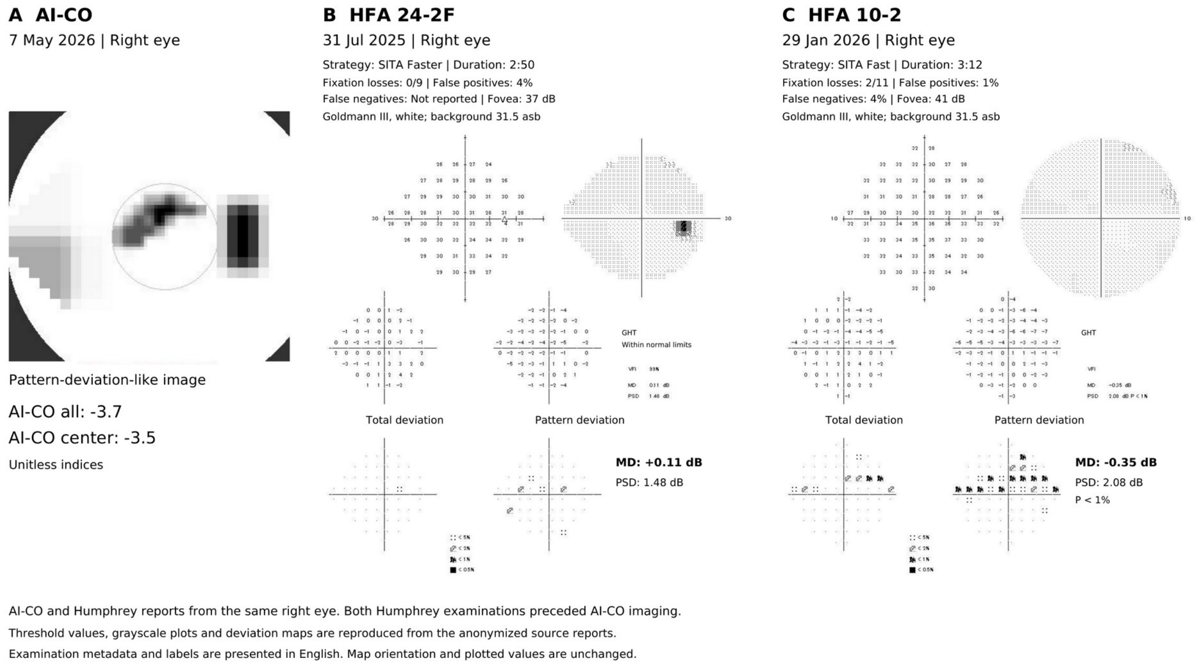


Supplementary Figure 2. AI-CO and Humphrey visual field reports from the same right eye. (A) AI-CO pattern-deviation-like image acquired on 7 May 2026; AI-CO all −3.7 and AI-CO center −3.5 are unitless indices. (B) Humphrey 24-2 SITA Faster examination performed on 31 July 2025 (MD +0.11 dB). (C) Humphrey 10-2 SITA Fast examination performed on 29 January 2026 (MD −0.35 dB). Examination dates differ; both Humphrey examinations preceded AI-CO imaging. The threshold values, grayscale plots, and deviation maps are reproduced from the anonymized source reports, with English labels and examination metadata. Map orientation and plotted values are preserved. This example does not demonstrate that AI-CO prompted additional perimetry.
